# Translation of Immunomodulatory Therapy to Treat Chronic Heart Failure: Preclinical Studies to First in Human

**DOI:** 10.1101/2022.08.19.22278416

**Authors:** H. David Humes, Keith D. Aaronson, Deborah Buffington, Hani N. Sabbah, Angela J. Westover, Lenar T. Yessayan, Balazs Szamosfalvi, Francis D. Pagani

**Affiliations:** Department of Internal Medicine, University of Michigan; Ann Arbor, Michigan; Innovative Biotherapies; Ann Arbor, Michigan; Department of Medicine; Henry Ford Hospital, Detroit, Michigan; Department of Cardiovascular Surgery; University of Michigan, Ann Arbor, Michigan

## Abstract

The objective of this study was to evaluate the use of an extracorporeal immunomodulatory device on the immune dysregulated state of chronic heart failure (CHF). Inflammation has been associated with progression and complications of CHF but no effective therapy has yet been identified to treat this dysregulated immunologic state. The selective cytopheretic device (SCD) provides extracorporeal autologous cell processing to lessen the inflammatory activity of circulating leukocytes of the innate immunologic system. SCD treatment in a canine model of systolic CHF diminished leukocyte inflammatory activity and enhanced cardiac performance as measured by left ventricular (LV) ejection fraction and myocardial contractility by stroke volume (SV) up to 4 weeks after treatment initiation. Translation of these observations in first in human, proof of concept clinical study was evaluated in a patient with severe systolic CHF ineligible for cardiac transplantation or left ventricular assist device (LVAD) due to renal insufficiency and right ventricular dysfunction. Six hour SCD treatments over 6 consecutive days resulted in selective removal of inflammatory neutrophils and monocytes and reduction in key plasma cytokines, including tumor necrosis factor (TNF)-a, interleukin (IL)-6, IL-8, monocyte chemoattractant protein (MCP)-1. These immunologic changes were associated with significant improvements in cardiac power output, right ventricular stroke work index, cardiac index and LVSV index, stabilization of renal function with progressive volume removal permitted successful LVAD implantation. This translational research study demonstrates a promising immunomodulatory approach to improve cardiac performance in systolic CHF and supports the important role of inflammation in the progression of CHF.

## INTRODUCTION

Chronic systolic heart failure (CHF), also referred to as chronic heart failure with reduced ejection fraction (HFrEF), is due primarily to the loss of left ventricular contractile function. Patients suffering from CHF have a poor prognosis with a 50% mortality rate within 5 years after initial diagnosis, despite pharmacologic and interventional therapies (1). Inflammation has been associated with the development, progression, and complication of HFrEF. Elevated levels of cytokines are found in plasma and myocardial tissue in CHF patients compared to normal controls (2.3). Proinflammatory cytokines, including tumor necrosis factor (TNF)-a and interleukin (IL)-6, diminish myocardial contractility (2,3). CHF patients have neutrophilia due to a higher percentage of neutrophils with delayed apoptotic progression as well as higher circulating absolute monocyte counts compared to healthy controls (4-6). In addition, high percentages of the proinflammatory intermediate subset of circulating monocytes (CD14^+^ CD16^+^) are associated with CHF progression (7,8). Translation of these observations to successful approaches to treat this chronic disease process has been disappointing, questioning the role of inflammation in the pathophysiology of HFrEF (9).

Despite growing evidence that acute and chronic inflammation promoted by neutrophil and monocyte/macrophage dysregulation is associated with CHF progression and poor outcome, an approach to immunomodulate the dysregulated leukocyte activity in CHF is currently an untested paradigm. The data presented in this report describe the evaluation of a novel immunomodulatory device, the selective cytopheretic device (SCD), on the immune dysregulated state of CHF and assess the potential benefit of this innovative strategy to improve the cardiovascular function in systolic CHF. The SCD is a polycarbonate cartridge containing hollow polysulfone membranes and is deployed in an extracorporeal blood circuit. This device preferentially binds activated circulating leukocytes (LE), primarily neutrophils and monocytes, in the low calcium environment afforded by regional citrate anticoagulation (RCA). These bound LE are deactivated and released back to the systemic circulation, resulting in a diminution of the dysregulated inflammatory states of acute or chronic organ dysfunction (10). This continuous autologous cell processing activity results in measurable diminution of excessive inflammatory responses with improvement of solid organ dysfunction in a variety of preclinical and clinical studies, including sepsis, acute kidney injury, ischemia/reperfusion injury, intracerebral hemorrhage, cardiopulmonary bypass, adult respiratory distress syndrome, chronic kidney disease, and type 2 diabetes mellitus (10-19).

This report provides evidence that SCD treatment in a well-established canine model of HFrEF improves myocardial contractility and left ventricular ejection fraction (LVEF) up to four weeks after treatment initiation. This preclinical observation allowed the translation of this approach to a first-in-human, proof of concept evaluation in a subject with longstanding biventricular failure to successfully bridge to left ventricular assist device (LVAD) implantation.

## MATERIALS AND METHODS

### Experimental Design

#### Canine Model

Male mongrel dogs weighing between 21 and 29 kg were used in these studies. CHF with HFrEF was induced in these animals using well established published procedures (20). Utilizing sterile techniques and cardiac catheterization, coronary microembolization using polystyrene microspheres (70-102 um in diameter) was accomplished to promote small left ventricular infarcts while the animals were under general anesthesia. Multiple sequential microembolizations were performed 2 weeks apart until the LVEF was less than 35% as determined by angiography. The animals were allowed to recover for at least 6 weeks after the last embolization before proceeding to the treatment protocol. All animal procedures and protocol were approved by Institutional Animal Care and Use Committee and conformed to the “Position of the American Heart Association on Research Animal Society”.

#### Initial Acute Canine Study (Citrate vs Heparin)

Five dogs with advanced chronic heart failure were evaluated with short term SCD treatment for 4 hours. Three animals were treated with SCD and RCA and two animals were treated with SCD with heparin anticoagulation. For these initial studies, SCD treatment is defined as SCD with RCA and sham control treatment is defined as SCD with heparin anticoagulation.

Access to the extracorporeal circuit, consisting of a single pump, single cartridge system, was accomplished by the insertion of a central venous double lumen catheter into the right jugular vein with connection to arterial and venous hemodialysis lines pre and post SCD (Figure 1). A prototype SCD with a membrane surface area of 1.4 m^2^ was used and blood flow was set at 120mL/min. For SCD-RCA studies, citrate (ACD-A) was given at the arterial outlet at 180mL/hr to maintain circuit iCa below 0.4 mmole/L and calcium chloride 2% solution was given at the inlet of the venous line at 50mL/hr to maintain systemic blood calcium at 0.9-1.4 mmol/L. Sham animals were systemically heparinized, and activated clotting time measured to ensure patency of the circuit. Animals were instrumented with a Swan-Ganz catheter, and hemodynamic measurements taken minimally at baseline, 5 min, 2, 4, 6 hours. Treatment was stopped at 4 hours and the 6-hour measurements were after a 2-hour washout period. Ventriculograms were recorded at baseline and after 4 hours of therapy. At the end of each 6-hour session, dogs were euthanized.

**Figure 1.**
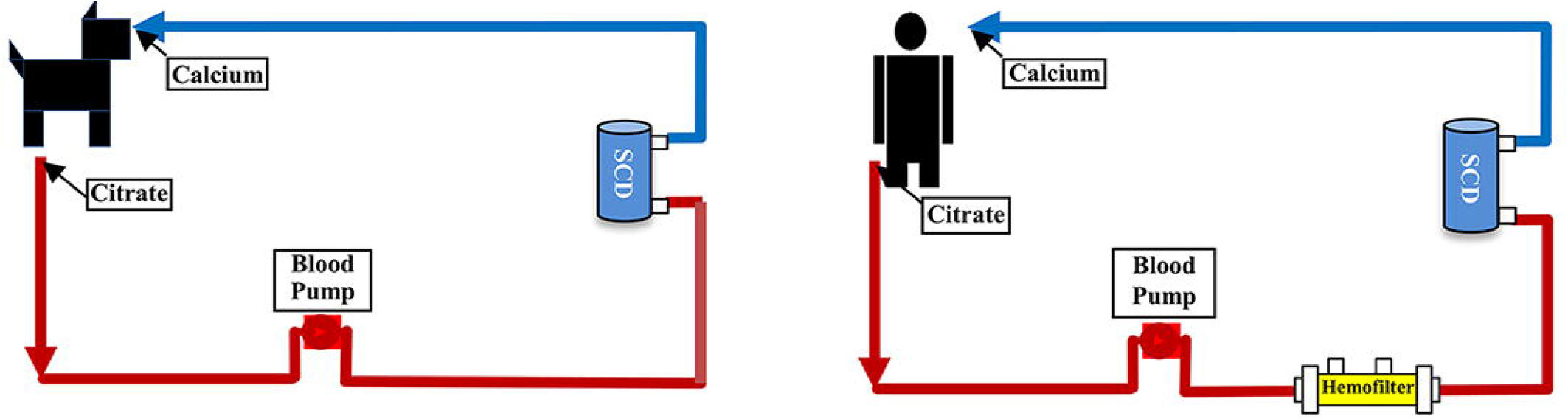
Schematic of extracorporeal blood circuits integrating SCD in canine and human studies. Left panel displays the extracorporeal circuit for SCD treatment in the canine model of CHF/HFrEF. Blood flow rate was 100ml/min, citrate (ACD-A) was administered at 180 ml/hour and 2% CaCl2 was given at 40 ml/min to maintain systemic and circuit iCa between 0.9 to 1.4 and 0.25 to 0.4 mmol/L, respectively. Right panel displays the exrcorporeal circuit for SCD treatment for human subjects. Blood flow rate was 80 ml/min. Citrate (ACD-A) and CaCl_2_ were administered as per protocol (Supplemental Appendix) to maintain systemic and circuit iCa between 0.9-1.4 and 0.25-0.4 mmol/L, respectively. The hemofilter was placed in the circuit to improve citrate removal to minimize any tendency to citrate toxicity during treatment.

#### Chronic Canine Studies (SCD vs SHAM)

Twelve dogs with advanced CHF (LVEF<35%) were used as follows: 5 dogs received sham treatments and 7 dogs received SCD treatments. For these studies, SCD treatment is defined as SCD with RCA and sham control treatment is defined as a RCA circuit only. The sham and SCD treatments were for 6 hours. SCD treatment was administered three times over one week with intervals of 48 or 72 hours between treatments in 5 CHF animals and once a week for three weeks in 2 animals. Initial baseline measurements were made at week 0 with cardiac catheterization and hemodynamic measurements. At the end of these measurements, blood was drawn and collected for inflammatory indices and serum analytes. Starting 48 hours after baseline assessments, three 6-hour treatment sessions of sham or SCD therapy (labelled S1, S2, S3) as described above. All animals were followed for 4 weeks after initiating the first SCD treatment. For analysis the data from the 7 SCD treated CHF animals were combined since the effect of SCD treatment on cardiovascular parameters were similar among all the treated animals. Specifically, the first 5 animals received 3 SCD treatments over 5-7 days and durable cardiac functional improvements were observed over the subsequent 3 weeks of observation. Accordingly, the next 2 animals were treated once weekly for 3 weeks and followed for an additional week for a 4 week observational period. The spacing of SCD treatment in 1 week intervals in animals was done to assess a less frequent treatment interval from 2 to 3 days to 7 days. Since the SCD treatment effects were similar in the first 5 animals compared to the last 2 animals, the data from the 7 (5+2) animals were combined for comparisons to the 5 untreated control animals.

Establishment of the extracorporeal circuit was accomplished as described above. A prototype SCD with a membrane surface area of 1.0 m^2^ was used and blood flow was set at 100 mL/min. Regional citrate anticoagulation of the circuit was performed with citrate (ACD-A) administered at 180 mL/hr at the outlet of the venous catheter and 2% CaCl_2_ given at 40 mL/hr at the inlet of the catheter. Systemic and circuit iCa were measured hourly to maintain levels between 0.9-1.4 and 0.25-0.4 mmol/L, respectively. Cardiac and hemodynamic parameters were re-evaluated at 48 hours, week 1, once either at the end of week 2 or beginning of week 3, and week 4 of this treatment and follow up period. Blood was collected at the end of each cardiac catheterization procedure.

Hemodynamic measurements were made during left and right heart catheterizations in anesthetized dogs using well established procedures (20). Left ventriculograms were performed during cardiac catheterization after completion of the hemodynamic measurements. Ventriculograms were performed with a pulse injection of 20 mL of contrast material (RENO-M-60, Squibb Diagnostics, New York, NY). At the end of the 4 week follow up period, dogs were euthanized.

#### Analysis of SCD associated cells_and assessment of Leukocyte cell surface markers of activation

At the end of each session, after returning blood from the blood circuit to the dog, the SCD cartridge was disconnected from the circuit after flushing with one liter of normal saline. The extra-capillary space of the cartridge was filled with a cell detachment solution consisting of 0.2% EDTA in normal saline for at least one hour. The eluted cells were then collected and analyzed (11).

#### Assessment of Leukocyte cell surface markers of activation

Levels of leukocyte cell surface markers were evaluated from both systemic blood and SCD membrane associated leukocytes (See Supplemental Appendix (Methods) for more detail). Complete blood counts were measured with a Hemavet 950 automated analyzer (Drew Scientific). Cytokine concentrations of IL-6 and TNF-a were measured with commercial enzyme linked immunosorbent assay (ELISA) kits reactive to canine cytokines (R&D Systems).

### Human Study

The first in human study using SCD treatment in a patient with HFrEF was undertaken with a FDA approved IDE G180055 entitled “Investigator Initiated Pilot Study to Assess the Safety and Efficacy of a Selective Cytopheretic Device (SCD) to Treat ICU Patients with Acute on Chronic Systolic Heart Failure with Cardiorenal Syndrome Awaiting Left Ventricular Assist Device Implantation” and with local IRB approval (clinicaltrials.gov NCT03836482). Inclusion and Exclusion criteria are detailed in Table 1. The SCD-1.0 and its associated bloodlines (SeaStar Medical, Denver, CO) were integrated into a CRRT blood circuit using a Prismaflex pump system and HF1000 hemofilter (Baxter, Deerfield, IL) as detailed in Figure 1. The SCD is in series with the hemofilter. The blood circuit was connected to a double lumen intravenous hemodialysis catheter placed in the jugular vein to achieve a blood flow rate of 80 mL/hr. SCD formulation and and general treatment implementation is detailed in Supplemental Appendix (Methods).

**Table 1.**
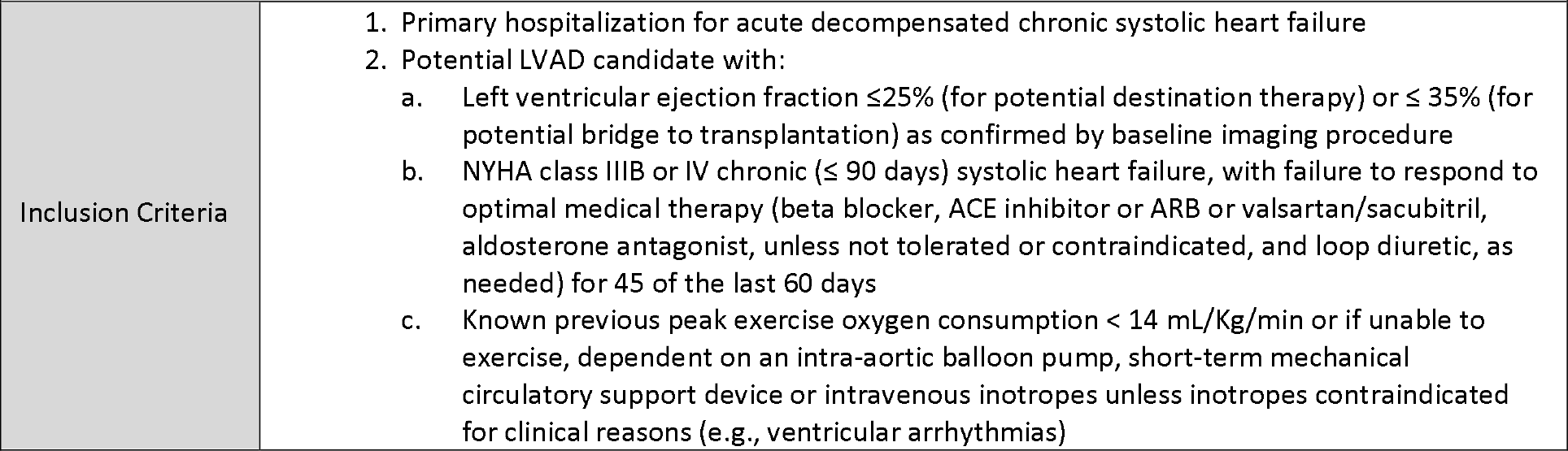
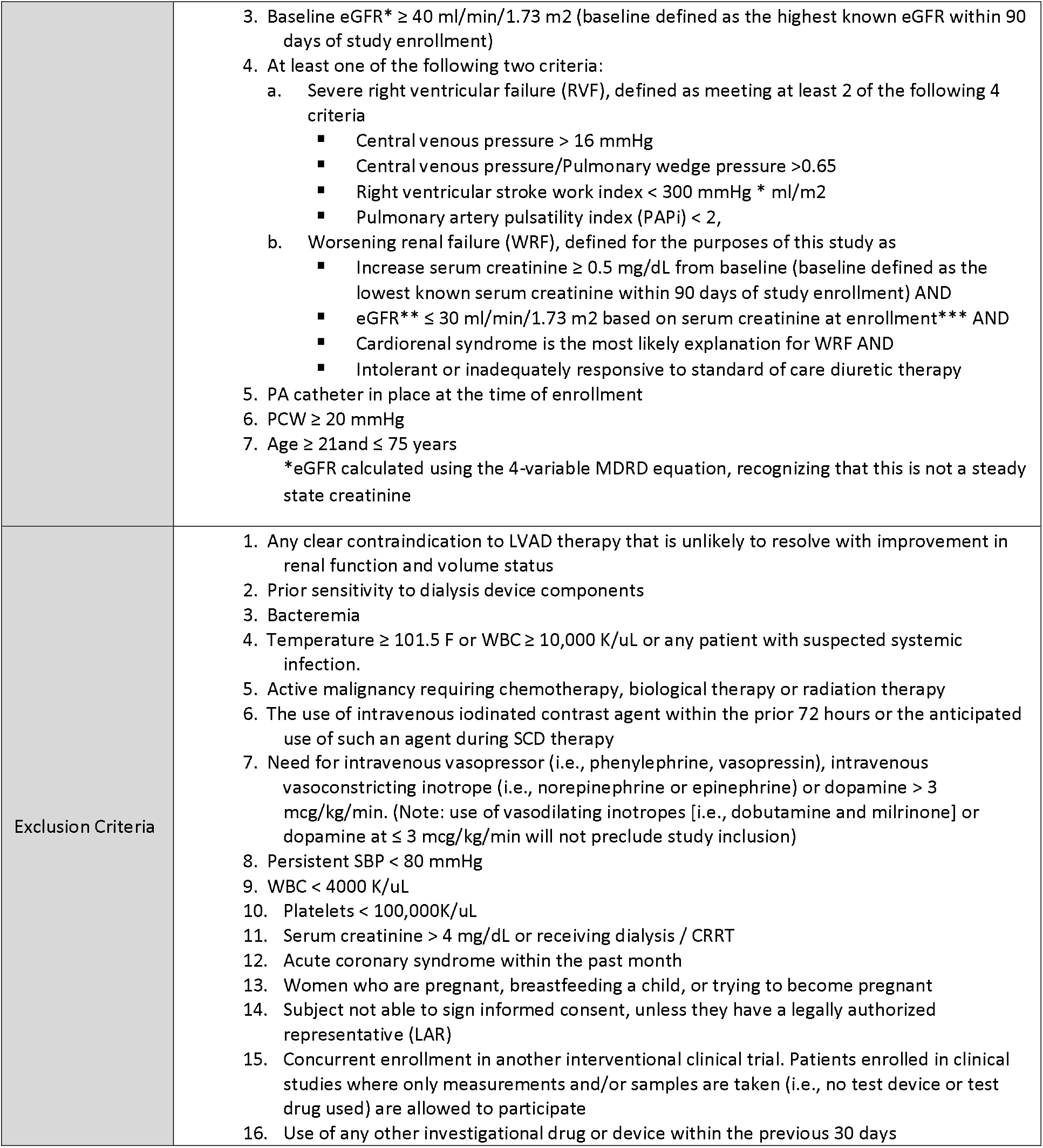
Inclusion and Exclusion Criteria of Clinical Study.

The patient underwent therapy according to clinical protocol with SCD treatment for 6 hours daily for 6 consecutive days at which time a decision to proceed to LVAD implantation was made. Standard of practice (SOP) clinical laboratory values and 24-hour collections of urine for volume and analytes were obtained daily. With the presence of a Swan-Ganz catheter, cardiac and hemodynamic parameters were measured according to SOP protocol and recorded. Research blood samples for cytokines, biomarkers and cytometric analysis were obtained daily just prior to SCD treatment.

#### Flow Cytometry and Cytokine Analysis

To correlate the clinical outcomes of SCD treatment and leukocyte parameters, flow cytometry was undertaken to see changes in cell surface markers of circulating and SCD bound neutrophils and monocytes before, during and after SCD treatment. Demonstration of SCD removal of activated leukocytes with changes in circulating phenotypes would link the SCD effects and immunologic rebalancing of the CHF associated dysregulated inflammatory state in CHF. An additional analysis was undertaken to evaluate whether SCD removal of substantive numbers of highly activated circulating leukocyte effector cells were able to diminish systemic levels of cytokines. Details of materials and methods for human leukocyte cytometric analysis and cytokine assays are included in Supplemental Appendix. Methods.

### Statistical Methods

All data are expressed as mean ± SE. Effects of SCD on various clinical and immunologic parameters within the treated group were evaluated with paired Student’s t test. Statistical comparisons between control and SCD treated groups were accomplished with analysis of variance (ANOVA) or non-paired t tests. Statistical significance was defined as p<0.05.

## RESULTS

### Preclinical Canine Model of CHF: SCD Treatment Effects on Cardiovascular Parameters

Five dogs with advanced chronic heart failure were evaluated with short term SCD treatment for 4 hours (Figure 1) Three animals were treated with SCD/RCA and two animals were treated with SCD/Heparin. As demonstrated in Figure 2, SCD/RCA treated animals increased their LVEF and stroke volume (SV) at four hours from baseline averages of 33.9±2.3 % and 26.7±4.9 ml to 46±4% and 35.3±7.3 ml, respectively, versus no change in SCD/Heparin treated animals averaging 32.8±2.3% and 26.0±1.0 ml to 34.0±0.3% and 25.5±1.5 ml. Ventriculograms in the SCD/RCA group demonstrated enhanced left ventricular contractility as a basis for the improved LVEF.

**Figure 2.**
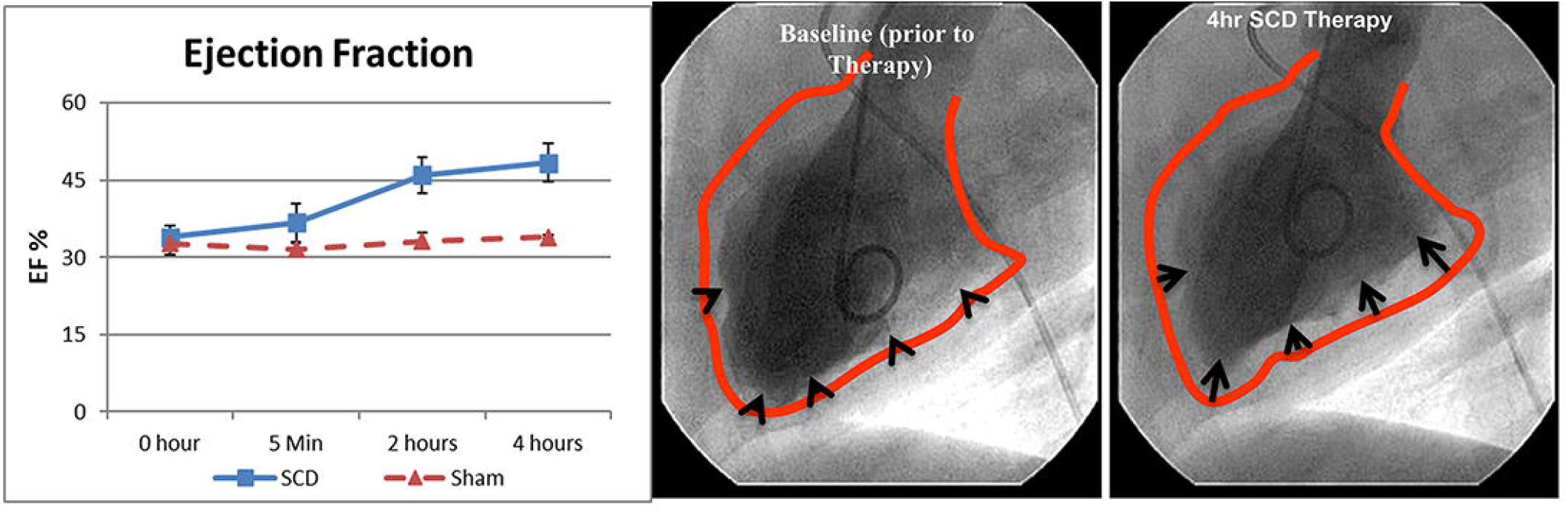
SCD treatment effects on cardiac performance in CHF dogs treated for 4h with SCD (n=3) or sham (n=2). LVEF (Left Panel) was returned near normal levels of 50-55% under SCD treatment. No effect on EF with sham therapy with systemic heparin anticoagulation was observed. Ventriculograms of a CHF dog heart (Right Panel) are shown at baseline (before therapy) and at the end of the 4h therapy session. The red line depicts the border of the diastolic silhouette overlayed on the systolic image, demonstrating improved contractility (black arrows) of the left ventricle after SCD treatment.

To extend these preliminary observations and to evaluate the durability of SCD treatment on this disease process, twelve additional dogs with well-established CHF (EF<35%) were used as follows: 5 dogs received sham treatments and 7 dogs received SCD treatments(see Methods).

Hemodynamic and angiographic measurements in normal dogs are displayed in Table 2 and have been reported previously (20). Both the control and treated groups after micro-embolization had evidence of chronic systolic heart failure with reductions in cardiac output (CO), LVEF and LV stroke volume (LVSV). During the 4-week evaluation period, no change in cardiac parameters compared to baseline values were observed in the sham group in contrast to the significant improvements observed in CO, LVEF, LVSV, and LV end systolic volume (LVESV) in the SCD treatment group. Mongrel dogs used for these studies had large variations in heart size due to breed and overall size of each animal. Therefore, the differences between treated and control groups were more readily apparent when the parameters were evaluated as values normalized to baseline. In this regard, SCD treatment increased CO, LVEF, LVSV, by greater than 15% (p<0.02, p<0.001, p<0.01, respectively) compared to sham treatment, as displayed in Figure 3.

**Table 2.** Cardiac Parameters in CHF Dogs.

| Control | Normal | Baseline | 48 Hr | 1-2 wk | 4wk |
| --- | --- | --- | --- | --- | --- |
| HR (beats/min) | 82±1 | 86±1 | 86±4 | 80±2 | 84±2 |
| MAP (mmHg) | 102±7 | 73±5 | 75±2 | 70±3 | 72±3 |
| SVR (dynes/seconds/cm-5) | 1810±110 | 3100±296 | 3254±160 | 3094±132 | 2982±191 |
| CO (L/min) | 4.35±0.7 | 1.92±0.07 | 1.92±0.09 | 1.84±0.04 | 1.94±0.08 |
| LVEF (%) | 59±3 | 34±2 | 34±2 | 35±2 | 34±2 |
| LVSV (ml) | 53±4 | 22±1 | 22±1 | 23±1 | 23±1 |
| LVESV (ml) | 24±2 | 44±4 | 43±5 | 43±5 | 44±5 |
| LVEDV (ml) | 60±7 | 66±4 | 65±5 | 66±5 | 67±5 |
| LVEDP (mmHg) | 6±1 | 13±0 | 14±1 | 14±0 | 15±0 |
| SCD Treatment | Normal | Baseline | 48 Hr | 1-2 wk | 4wk |
| HR (beats/min) | 82±1 | 85±1 | 85±4 | 85±2 | 85±2 |
| MAP (mmHg) | 102±7 | 74±3 | 81±4 | 82±4 | 77±4 |
| SVR (dynes/seconds/cm-5) | 1810±110 | 3361±150 | 3533±308 | 3458±222 | 3199±139** |
| CO (L/min) | 4.35±0.7 | 1.67±0.06 | 1.90±0.18 | 1.91±0.06*** | 1.94±0.12* |
| LVEF (%) | 59±3 | 34±1 | 39±1** | 39±1*** | 39±2** |
| LVSV (ml) | 53±4 | 20±1 | 22±1* | 23±1*** | 23±1** |
| LVESV (ml) | 24±2 | 38±2 | 35±2** | 35±2*** | 35±2** |
| LVEDV (ml) | 60±7 | 58±2 | 58±2 | 58±2 | 58±2 |
| LVEDP (mmHg) | 6±1 | 14±1 | 15±1 | 14±1 | 13±1 |

Values presented as mean ± SE.
Compared to Baseline, \*P<0.05, \*\*P<0.01, \*\*\*P<0.001.
HR=Heart Rate, MAP=Mean Arterial Pressure, SVR=Systemic Vascular Resistance, CO=Cardiac Output,
LVEF=Left Ventricular Ejection Fraction, LVSV=Left Ventricular Stroke Volume, LVESV=Left Ventricular
End-Systolic Volume, LVEDV=Left Ventricular End-Diastolic Volume, LVEDP=Left Ventricular End-Diastolic
Press

**Figure 3.**
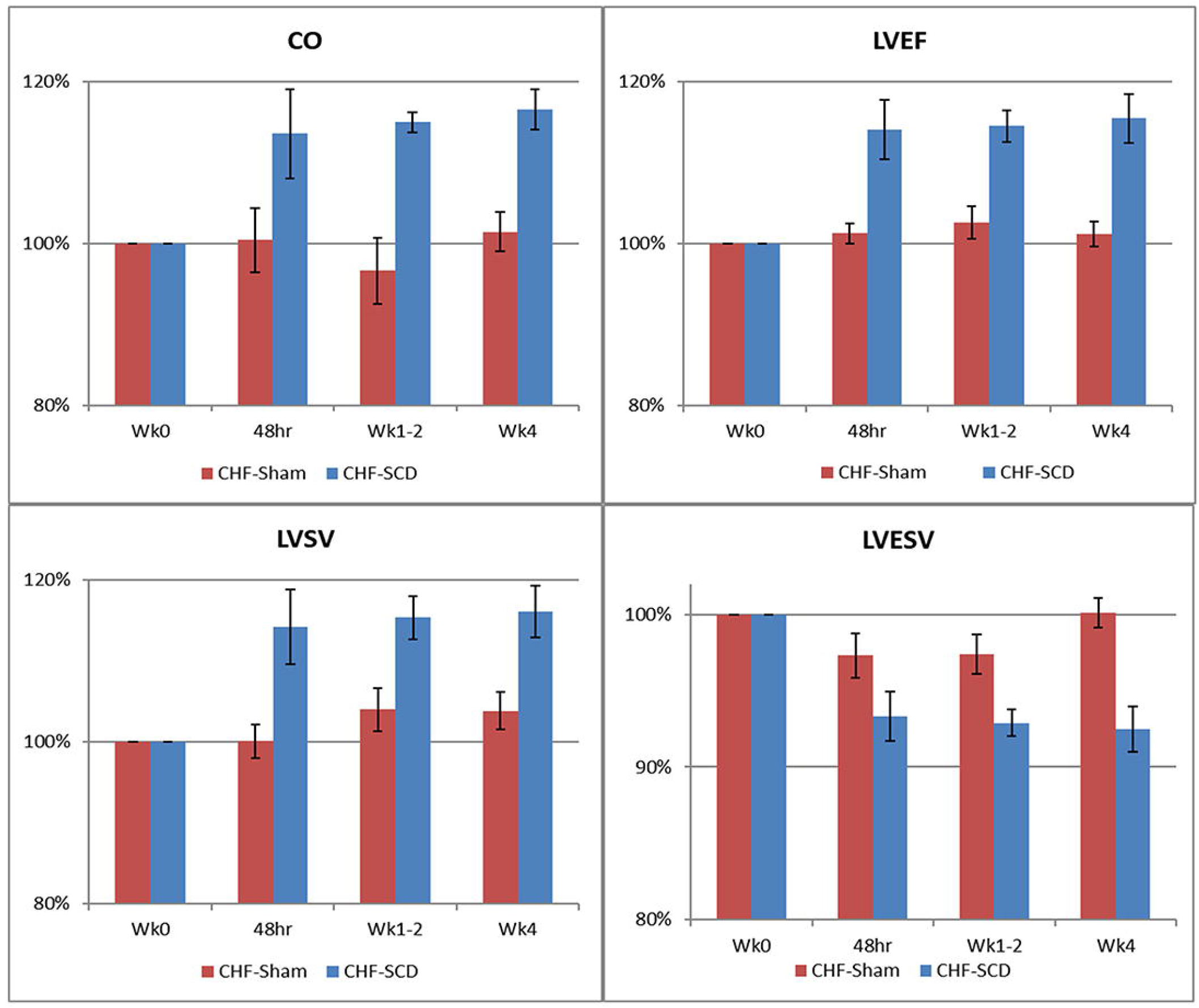
SCD treatment improves cardiac parameters compared to sham controls. SCD treatment (CHF-SCD) significantly increased cardiac output (CO), left ventricular (LE) ejection fraction (EF), LV stroke volume (SV), and decreased LV end systolic volume (ESV) compared to sham treatment (CHF-Sham) by ANOVA repeated measures over a 4 week time course; p<0.02, p<0.001, p<0.01, p<0.001, respectively.

### Preclinical Canine Model of CHF: SCD Treatment Effects on Immunologic Parameters

At the end of each treatment period (S1, S2, S3), cells bound to the SCD were eluted from the cartridge and analyzed. For the 3 SCD treatment periods, on average, 1.17 ± 0.34 × 10^9^ leukocytes were eluted from the SCD with 84 ± 1% neutrophils, 9±1% monocytes, and 7±1% lymphocytes and eosinophils, representing 6% and 5% of the circulating pool of neutrophils and monocytes, respectively. Flow cytometric measurement of the mean fluorescent intensity (MFI) of cells labeled with fluorochrome conjugated antibodies to targeted epitopes provides a relative measure of surface expression. The MFI of membrane associated CD11b labeled neutrophils was 11.5x higher than MFI of circulating neutrophils (p<0.0002). The MFI of CD11b and CD14 labeled monocytes associated with the SCD membrane were 11.3 x and 1.58 x higher than circulating monocytes (p<0.0001 and p<0.003, respectively). These results demonstrate that the SCD bound the more activated circulating leukocytes. The capture of these cells within the SCD resulted in lower CD11b MFI of circulating neutrophils and lower CD14 MFI in circulating monocytes during the 4-week time course of the study, as displayed in Figure 4. Circulating neutrophil surface expression of CD11b was consistently lower in SCD treated compared to the sham group. Circulating monocyte CD14 surface expression were also significantly lower (p=0.03) in the SCD group. Higher rates of neutrophil apoptosis were seen in the bound cells compared to circulating neutrophils. Of the eluded neutrophils 50 ± 7 % were apoptotic after 24 hours while only 25 ±7% of the comparative circulating neutrophils were apoptotic. This observation suggests that a higher percentage of the bound neutrophils are progressing to apoptosis compared to the circulating pool. In addition, since the neutrophils that bind to the SCD are more activated, an even greater percentage of the originally bound neutrophils are in a delayed apoptotic state with an even lower percentage than 25% of the circulating pool.

**Figure 4.**
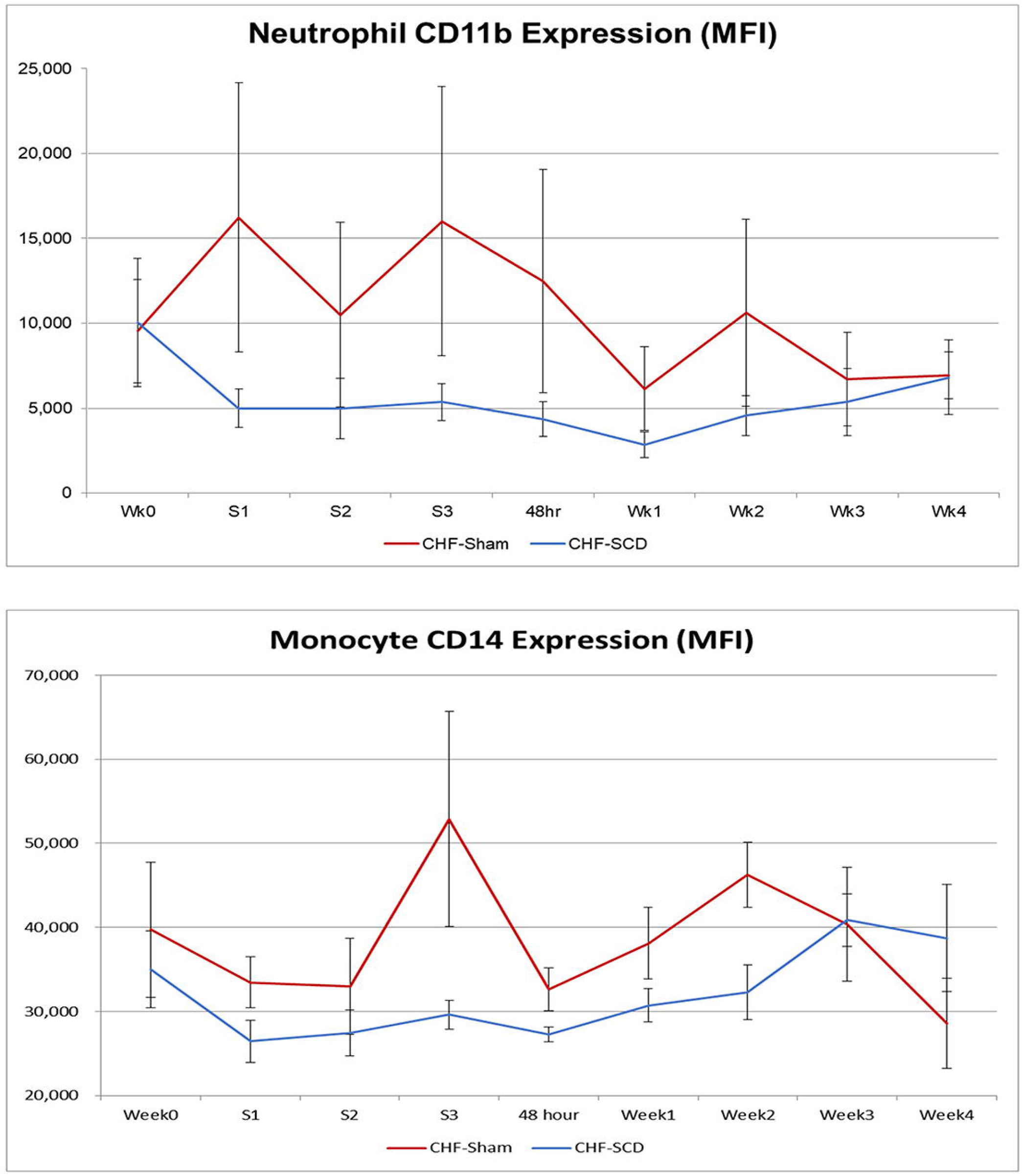
SCD treatments diminishes activation markers in circulating leukocytes compared to sham controls. SCD treatment lowered the MFIs of the cell surface markers CD11b for circulating neutrophils and CD14 for circulating monocytes compared to sham treatment at various time points during the 4 week evaluation period.

Interleukin (IL) -6 serum levels in SCD treated animals were significantly (p<0.02) lower than sham controls, averaging 4.10 ± 0.47 versus 10.33 ± 0.93 pg/mL, respectively, throughout the 4-week period. TNF-a levels did not differ between the two groups.

### First in Human. Proof of Concept

#### Medical Course

The subject was a male in his early 70s with longstanding slowly progressive idiopathic cardiomyopathy since 2000. In 2013 he had a LVEF of 20%. He was hospitalized at University Hospital 3 weeks prior with a 20-pound weight gain and increasing congestive symptoms. At that time, he was on an outpatient regimen of simvastatin, aspirin, sacubitril-valsartan, and beta-blocker. He obtained a right heart catheterization demonstrating a right atrial pressure (RAP) of 17 mmHg, pulmonary capillary wedge pressure of 38 mmHg and a cardiac index (CI) of 1.69. His renal function parameters were blood urea nitrogen (BUN) 42 mg/dL and serum creatinine (Scr) 1.74 mg/dL. He was treated with intravenous furosemide and metolazone with a subsequent 15-pound weight loss. He was evaluated for heart transplantation but was disqualified due to age and co-morbidities.

He was discharged but quickly gained 18 pounds and sought a second opinion at another medical center for transplantation or LVAD implantation. He again was disqualified for transplantation or LVAD due to age, decreased renal function and moderate right ventricular (RV) dysfunction. He was subsequently transferred back to University Hospital to optimize his cardiac hemodynamics with indwelling pulmonary catheter monitoring and aggressive intravenous administration of high dose diuretics and inotropic agents. His admitting laboratory blood values were WBC 5,200, Hgb 9.9 g/dL, Sodium 135 mEq/L, CO2 27 mEq/L, BUN 39 mg/dL, and Scr 2.63 mg/dL, BNP 1124 pg/mL. Echocardiogram showed RV systolic dysfunction and LVEF 10%. Treatment over the next 7 days with intravenous furosemide (40 mg/hr) and milrinone (0.3 µg/kg/min) resulted in a net fluid loss of 6.9 liters without improvement of RAP and modest increase in his cardiac index compared to values on admission to the ICU. His BUN and Scr increased to 57 and 2.69 mg/dL, respectively.

Due to lack of improvement in his right atrial pressure or renal function parameters, he was enrolled after meeting all clinical criteria for SCD treatment as a potential bridge for LVAD implanatation (IDE 180055; IRB approved; clinicaltrials.gov NCT038364482). After completing written informed consent, he was enrolled into the clinical trial. As part of this consent, he agreed to have the results of the research study to be published in medical literature. Accordingly, SCD therapy was initiated on Day 8 of his hospitalization. He was treated with SCD for 6 hours daily for 6 consecutive days. Also per protocol, no net volume removal with ultrafiltration or dialysis occurred during these 6-hour treatment periods or at any other times during the 6-day study.

#### Assessment of Cardiac Function

His cardiac parameters improved and demonstrated sustained improvement for the 6-day study period. As demonstrated in Table 3 and Figure 5, comparing daily cardiac parameters during the 6 days prior to SCD intervention to those during the 6 days of SCD treatment, significant improvements were observed: CO and cardiac index (p=0.023), LVSV and LVSV index (p=0.0009), right ventricular stroke volume index (RVSVI, p=0.008) and cardiac power output (CPO, p=0.017). During this 6-day period, he was continued on diuretic therapy and milrinone with a further net fluid loss of 5.7 L without worsening renal function parameters, Scr ranging from 2.5 to 2.81 mg/dL and BUN from 48-53 mg/dL. No serious adverse events with SCD treatment were observed. With his improved cardiac parameters and stable renal function, he underwent LVAD implantation 3 days after discontinuing SCD treatment. Of note, during those three days his Scr further improved to 2.34 mg/dL. After LVAD placement his Scr continued to decline over the course of 2 weeks to 1.48 mg/dL. He was subsequently discharged from the hospital without complications.

**Table 3.**
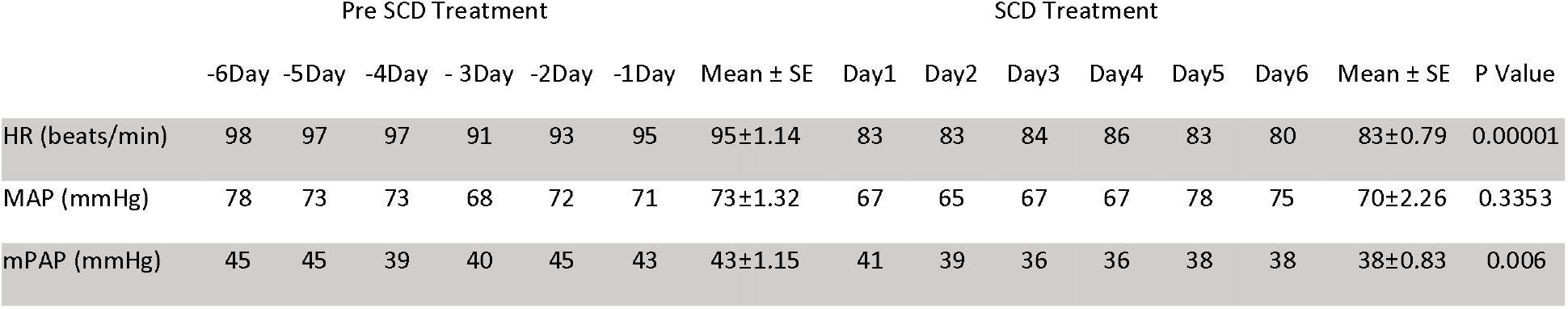
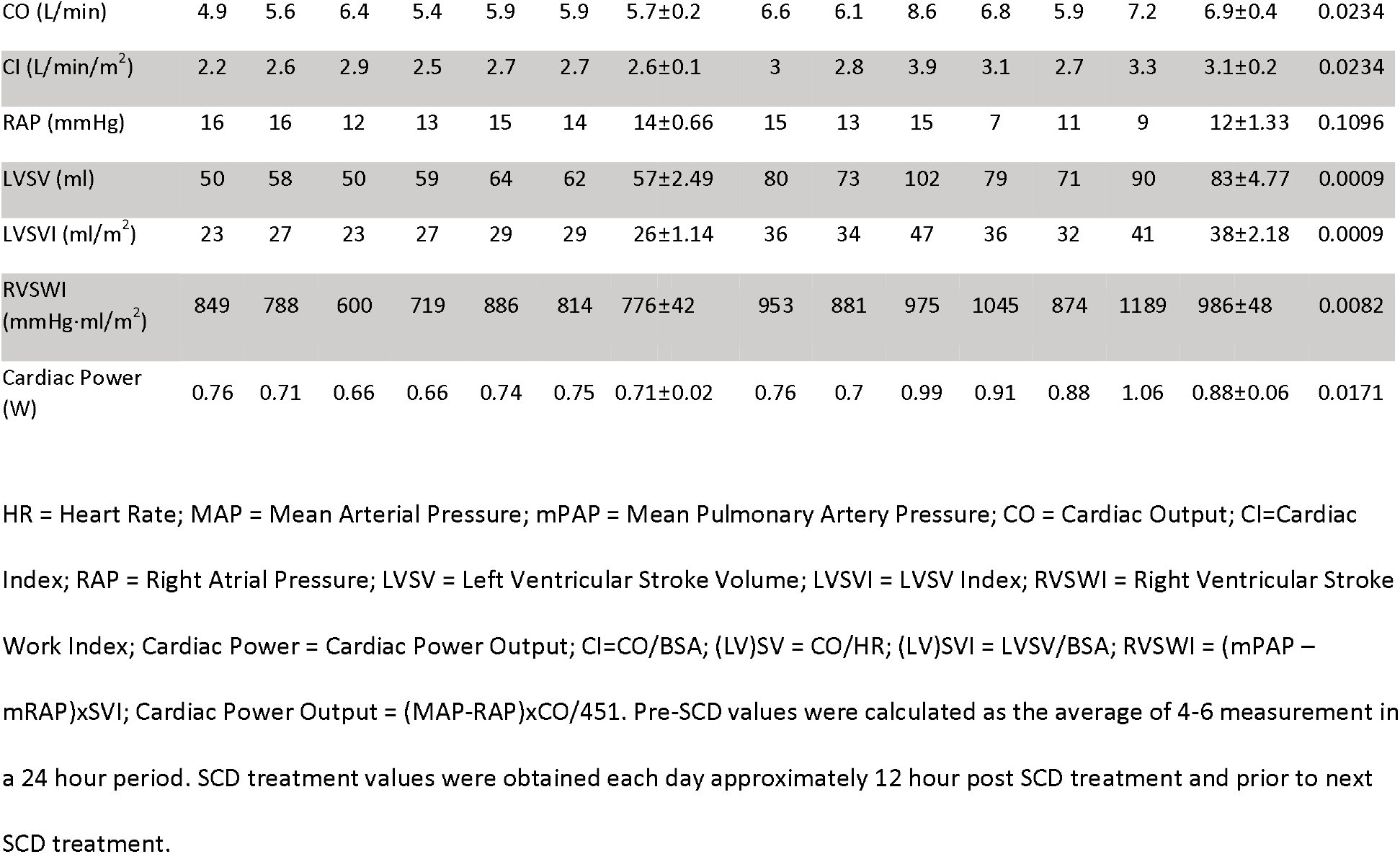
Cardiac Parameters of Enrolled Subject.

**Figure 5.**
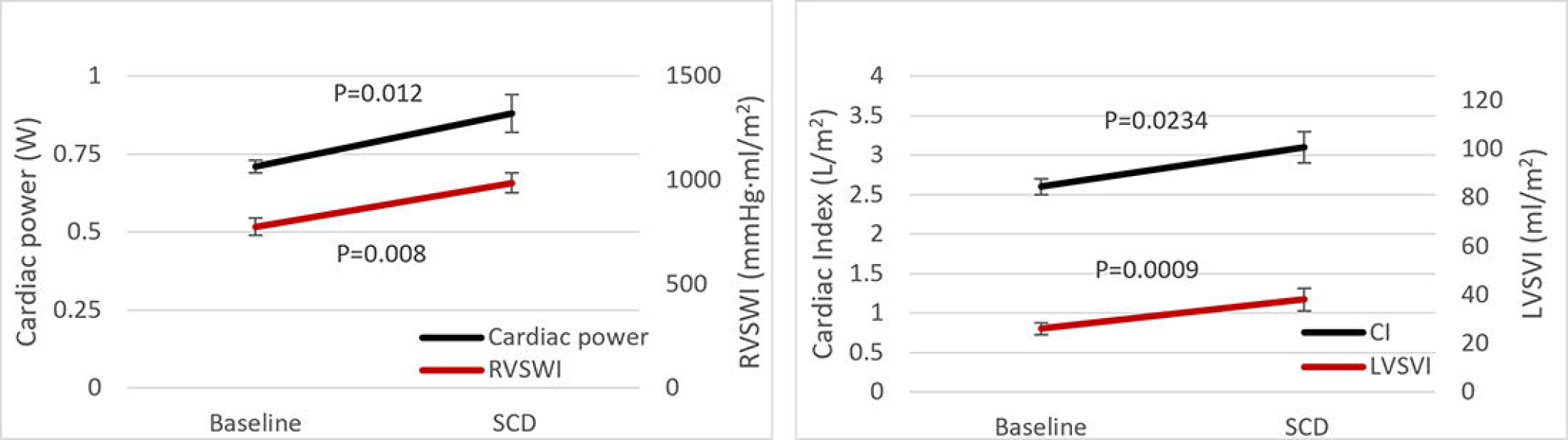
Effect of SCD Treatment on Cardiac Parameters in Enrolled Subject. Left panel. Improvements in Cardiac Power Output (CPO) and Right Ventricle Stroke Work Index (RVSWI) from Baseline (pre-SCD treatment, Day-6 to Day-1) and during SCD treatment (Day1-6). Right panel. Improvements in Cardiac Index (CI) and Left Ventricle Stroke Volume Index from Baseline and during SCD treatment.

#### Immunologic Assessment

Prior to, during and after SCD treatment, research samples were evaluated, as per IRB approval, to assess immunologic parameters and leukocyte cytometric analysis. As shown in Table 4, the patient was inflamed with elevated plasma levels of interleukin (IL)-6 and monocyte chemoattractant protein (MCP)-1. SCD treatment substantially decreased all measured plasma levels of cytokines: IL-6, MCP-1, IL-8, IL-10 and tumor necrosis factor (TNF)-a after only two days of SCD treatment compared to baseline values. Elution of the post treatment SCDs on days 1, 3, and 5 demonstrated 1.1 × 10^10^ (87% NE, 12% MO), 1.28 × 10^9^ (73% NE, 25% MO), and 8.29 × 10^8^ (72% NE, 28% MO), cells, respectively, were bound to the devices.

**Table 4.**
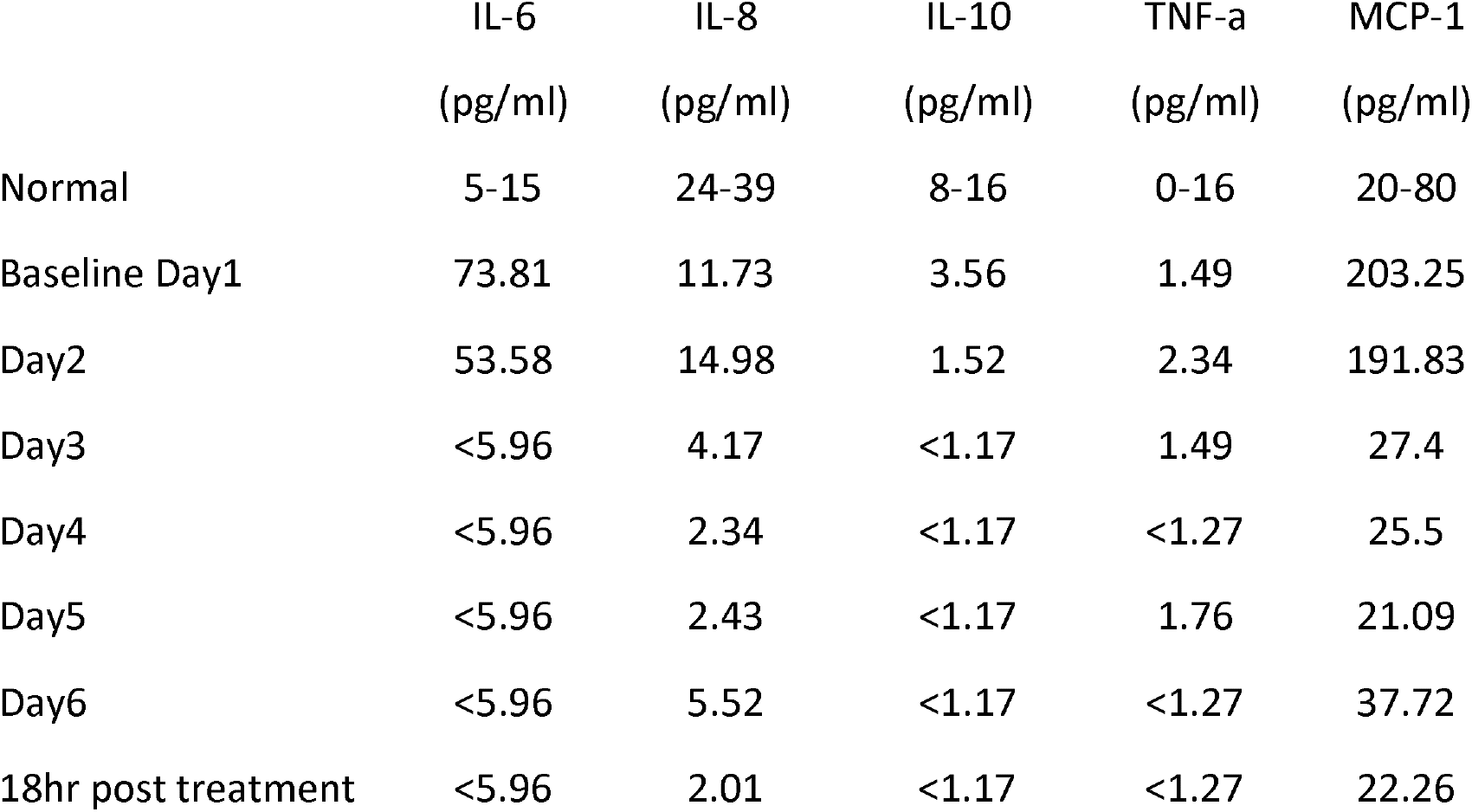
Serum Cytokine Levels and SCD treatment.

|  | IL-6 | IL-8 | IL-10 | TNF-a | MCP-1 |
| --- | --- | --- | --- | --- | --- |
|  | (pg/ml) | (pg/ml) | (pg/ml) | (pg/ml) | (pg/ml) |
| Normal | 5-15 | 24-39 | 8-16 | 0-16 | 20-80 |
| Baseline Day1 | 73.81 | 11.73 | 3.56 | 1.49 | 203.25 |
| Day2 | 53.58 | 14.98 | 1.52 | 2.34 | 191.83 |
| Day3 | <5.96 | 4.17 | <1.17 | 1.49 | 27.4 |
| Day4 | <5.96 | 2.34 | <1.17 | <1.27 | 25.5 |
| Day5 | <5.96 | 2.43 | <1.17 | 1.76 | 21.09 |
| Day6 | <5.96 | 5.52 | <1.17 | <1.27 | 37.72 |
| 18hr post treatment | <5.96 | 2.01 | <1.17 | <1.27 | 22.26 |

For cytometric analysis, two antibody panels were used, one to evaluate neutrophil activation and life cycle and a second for monocyte classification including CD14, CD16 and HLADR (Supplemental Appendix. Materials). As seen in Figure 6, cytometric analysis demonstrated that the SCD bound the more activated, mature circulating neutrophils. This observation was made due to the higher cell surface expression of CD11b and CD10 of 2.89 and 1.63-fold (p<0.03 and p< 0.04), as measured by MFI respectively, of SCD membrane bound cells compared to circulating neutrophils. The dramatic decline in the surface expression of CD62L (L-selectin) of the SCD associated neutrophils compared to circulating cells (p<0.0001) reflected the binding events occurring on the SCD membranes. L-selectin is shed from neutrophils upon attachment to endothelium and other surfaces (21). SCD also bound the monocytes with a higher surface expression of CD11b and CD14 with MFIs for these markers of SCD associated monocytes of 2.60 and 1.80-fold (p<0.004 and p<0.0005) higher than circulating monocytes, respectively.

**Figure 6.**
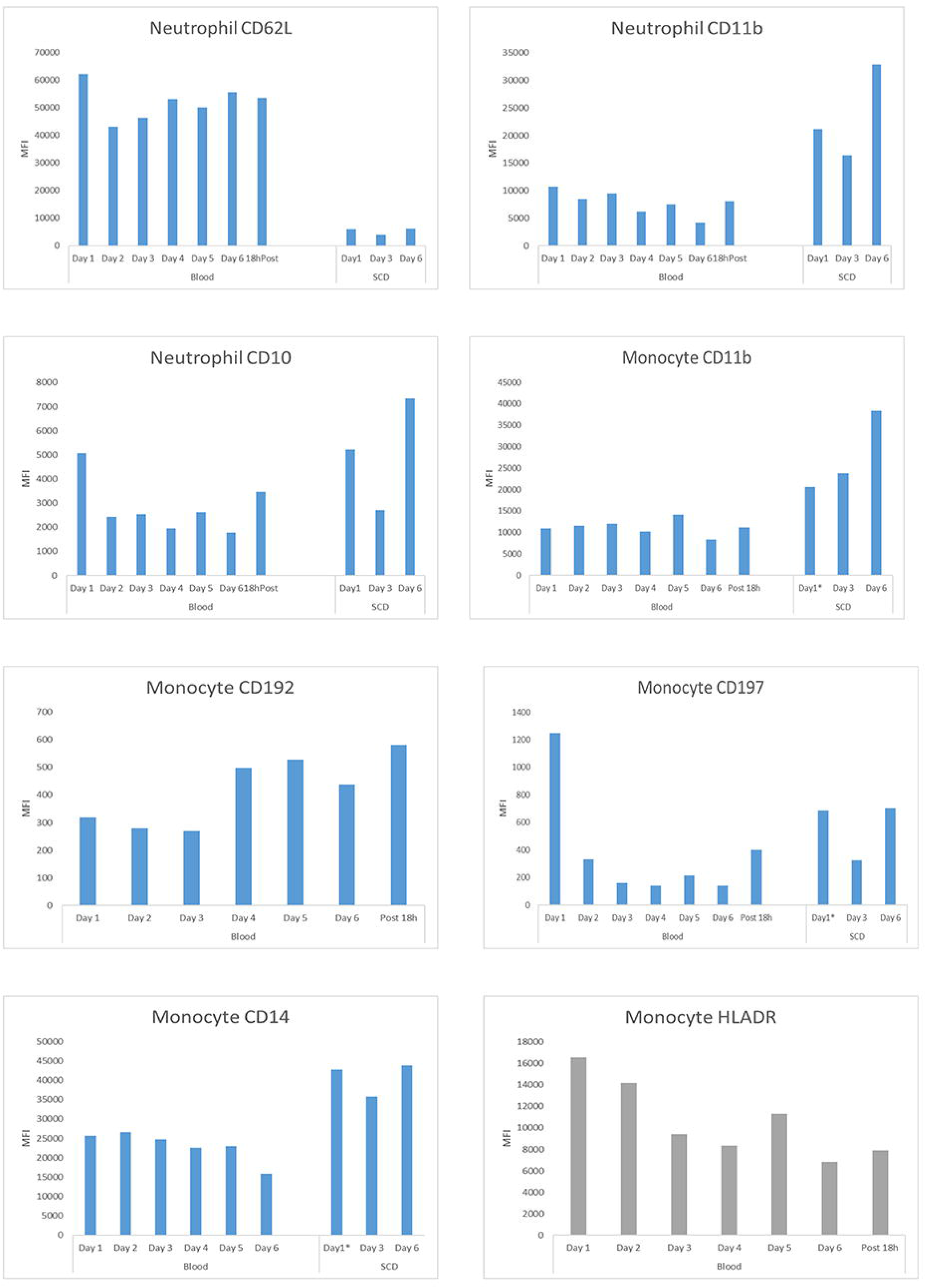
Effect of SCD treatment on leukocyte phenotypes in a patient with severe systolic CHF. Each graph displays the MFI of various cell surface markers on either circulating blood neutrophils or monocytes during the 6 day course of daily 6 hours of SCD treatment. Also displayed are the MFIs of the eluted neutrophils and monocytes from the SCD after treatments on day 1, 3, and 5. All monocyte graphs depict the MFI of the entire monocyte population except for the Monocyte HLADR graph which presents the MFI of the surface marker of HLADR in the intermediate (CD14+CD16+) monocyte subpopulation. Day 1 values were baseline measurements prior to initiation of SCD treatment. Day 2 and all subsequent Days represent values obtained during the morning after the prior day’s treatment.

To assess key cell surface markers for monocyte adhesion and migration into tissue, CD197 and CD192 were also analyzed. The MFI of circulating CD197 labeled monocytes decreased on average 6.3-fold compared to pre-treatment baseline levels with a 1.26-fold increase (p< 0.01) in the MFI of SCD bound CD197 labeled monocytes; whereas the MFI of circulating CD192 labeled monocytes increased after 3 days of treatment compared to baseline and prior levels on days 1-3 (p<0.002). HLA-DR labeled intermediate monocytes (gated using CD14^+^ CD16^+^) progressively declined during treatment. Table 5 displays the percentage of circulating and SCD bound classical, intermediate, and non-classical monocyte subsets before, during and after SCD treatment. Following the first two SCD treatments, there was a shift in the distribution of circulating monocytes away from the classical phenotype toward the intermediate phenotype with the classical monocyte subset constituting 77.3%, 60.4% and 67.3% and the intermediate monocyte subset comprising 11.6%, 29.5% and 18.5% of circulating monocytes at baseline, after Day 1 and after Day 2, respectively. The percentage distribution of circulating monocytes then reverted back by day 5 of treatment to the baseline distribution of 77.4, 11.6, and 10.6 % of classical, intermediate and non-classical phenotypes, respectively.

**Table 5.**
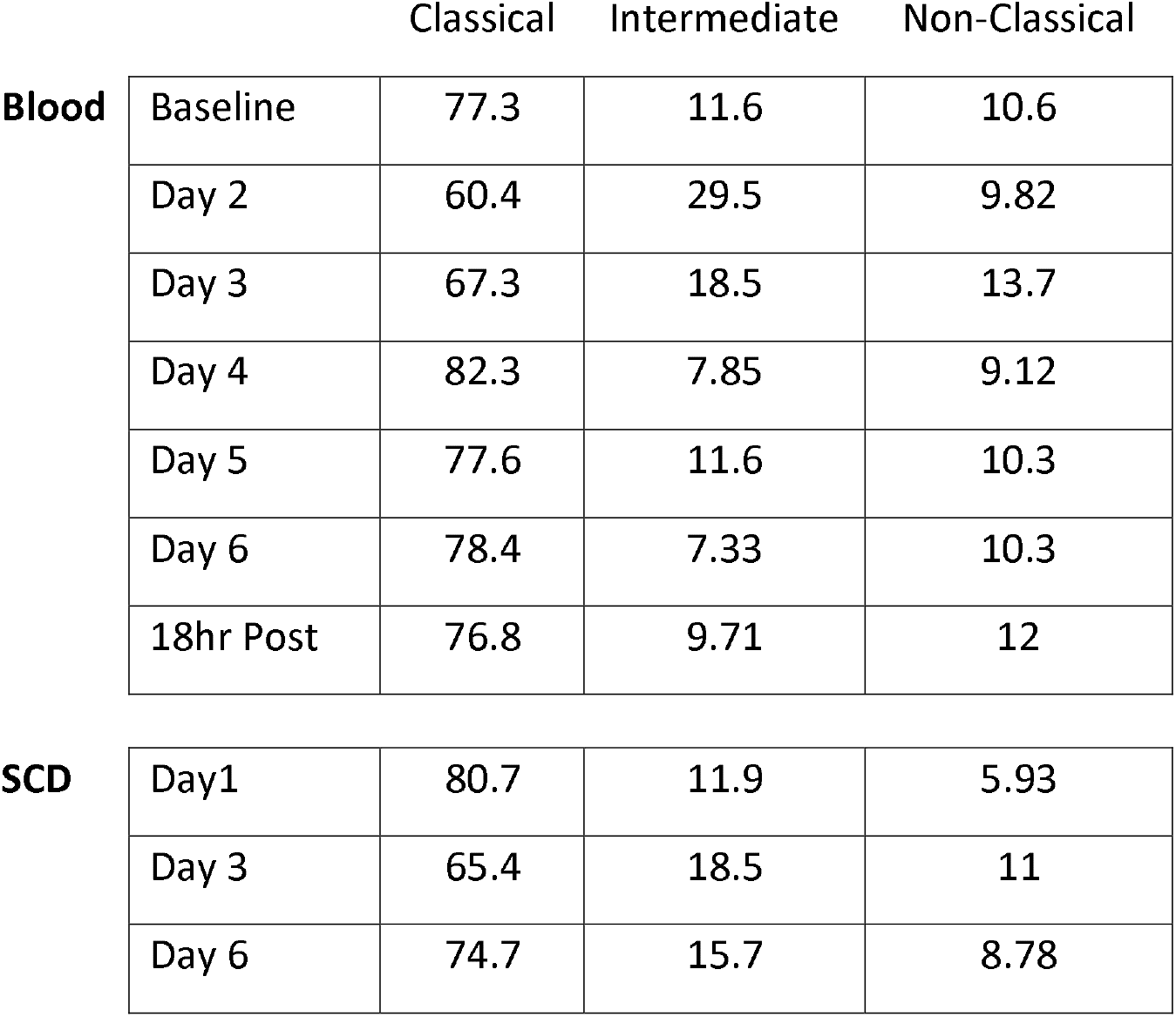
Monocyte Subsets in Enrolled Patient.

The elution of membrane associated cells from the SCD after the first, third and fifth days of treatment demonstrated that the SCD bound 29.9, 4.9, and 2.7% of the circulating pool of neutrophils and 21.6, 5.9, and 4.9% of the circulating pool of monocytes, respectively.

## DISCUSSION

CHF is associated with a chronic inflammatory disease state, especially related to the innate immunologic system, with increased activation of circulating neutrophils and monocytes (2,3,22), This inflammatory dysregulation may contribute to cardiac dysfunction. Strategies to reduce the cardio-depressant effects of acute and chronic inflammation in HFrEF have not yielded successful new therapies. Accordingly, the extracorporeal immunomodulation approach with SCD treatment warranted an evaluation first in a preclinical model of systolic CHF prior to translation into the clinical setting. In preclinical models, SCD therapy has shown efficacy in acute multiorgan injury in severe sepsis, cardiopulmonary bypass, intracerebral hemorrhage (ICH), and ischemia/reperfusion injury (IRI)(10-13,17). This device has been tested in 6 clinical studies in ICU patients with acute kidney injury and multiorgan dysfunction requiring dialysis and COVID-19 patients with adult respiratory distress syndrome (ARDS) with improved clinical outcomes and no device-related serious adverse events (14-16,18,19, 23).

This innovative immunomodulatory approach to HFrEF was considered due to a key role that the innate immunologic system may play in the acute and chronic myocardial injury resulting in progression of CHF. A vigorous inflammatory response occurs immediately after reperfusion to the ischemic myocardium since various molecular signals are generated by injured endothelial cells and cardiomyocytes (2). This response is eventually important in the wound healing and remodeling necessary to reestablish cardiac performance but is excessive and maladaptive. The increase in circulating levels of innate immune cells observed in CHF, including neutrophils and monocytes, arise both from the splenic monocyte reservoir pool and the bone marrow precursor pool to produce the initial pro-inflammatory response (24). The magnitude of neutrophil infiltration into the damaged area of the heart accentuates the degree of injury and cardiac dysfunction (25,26). The role of the circulating monocyte and its transformation into a tissue macrophage is now acknowledged to be central to the injury and repair phases of this process (27,28) A balanced monocyte/macrophage response, both in phenotype and timing, is necessary for optimal repair and healing (24,28). The suppression of the early phase of monocyte release from spleen reduces infarct size and myocardial dysfunction acutely and sub-acutely (29,30). In addition, more vigorous response of circulating monocyte proinflammatory phenotype after AMI has been correlated to a greater decline in left ventricular ejection fraction 6 months after AMI (31). The cardio-depressant effects of this immunologic activation have been well characterized (32-36). Chronic inflammation promoted by monocyte/macrophage dysregulation has been correlated with CHF progression and poor outcome and its suppression with mineralocorticoid receptor (MR) antagonists retard the progression and mortality in CHF patients (36,37). The modulation of this persistent inflammatory state associated with acute and chronic cardiac injury with SCD treatment may be an innovative approach to treat CHF and is the basis of this report.

Initial canine studies were remarkable considering it was unknown whether immunomodulating effects would be observed in this model or persist after therapy discontinuation. Changes were evident in all treated animals during acute treatment and through 2-hour washout period. Prior to continuing with canine studies, experience was obtained with another model of chronic inflammation, specifically metabolic syndrome in Ossabaw pigs. In this model, the greatest effects were observed when therapy was given in three sessions over 1 week and persisted for up to two weeks, indicating longer durability of treatment effects (13). Accordingly, the treatment protocol for the second set of canine studies was planned for three therapy sessions using intervals of 48 hours to one week and followed for 4 weeks after treatment initiation. In these preclinical experiments SCD treatment demonstrated significant improvements in CO, LVEF, LVSV, LVEDV compared to the baseline values as well as to the repeated measures of the sham control group over the entire 4-week observation period. These improvements in myocardial performance with SCD treatment were durable during the 4 weeks of observation.

In these CHF animals, cytometric analysis demonstrated that the SCD had sequestered 5-6% of circulating more activated leukocyte pools as assessed with cell surface markers of activation (CD11b, CD14, 38-40). The sequestration of these leukocytes was associated with declines in the inflammatory activity compared to sham controls of circulating neutrophils and monocytes in the treated animals (Figure 4). The reductions of these inflammatory leukocyte activation markers were accompanied with a significant reduction in serum IL-6 levels during the entire 4-week period, providing evidence of immunomodulation to a lessened inflammatory state. This immunomodulation results in improved cardiac performance both from a neutrophil effect to reduce systemic proinflammatory mediators and a monocyte effect to alter monocyte /macrophage trafficking into myocardial tissue with a less inflammatory phenotype (41,42).

With these encouraging findings in the preclinical studies, translation of this extracorporeal immunomodulation therapy into the clinical arena appeared to be worthy of evaluation. In this regard, a clinical protocol providing a compelling benefit to risk ratio was formulated to test this approach in a first in human, proof of concept.

Without heart transplantation or mechanical circulatory support, hospitalized individuals with Stage D acute on chronic systolic heart failure have a life expectancy of days to weeks. For refractory patients who are not eligible for heart transplantation, LVAD implantation is the only remaining treatment option. Due to high risks and poor outcomes, however, patients with poor renal function (eGFR < 30 mL/min/1.73m^2^) or right ventricular systolic heart failure (RVF) are excluded from LVAD candidacy at most centers. A clinical protocol was designed to evaluate whether SCD treatment in this clinical situation could improve renal function and/or right ventricular failure, an enrolled subject may be deemed eligible for LVAD implantation and proceed to a life sustaining procedure.

Accordingly, we enrolled our first patient who had evidence of a chronic inflammatory state and met all inclusion/exclusion criteria (See Supplemental Table S1). After patient consent, he was treated with SCD according to protocol. With SCD treatment, multiple cardiac parameters improved. Most notably, global cardiac performance and right ventricular contractility, as measured with CPO and RVSWI, were significantly increased. CPO is a strong predictor of outcome in patients with advanced CHF and preoperative RVSWI is also predictive of both RV failure and death post LVAD (43-46). With his improvement in RVSWI and stable renal function, he underwent successful LVAD implantation.

To assess the immunologic changes during SCD treatment, cell-sorting and cytometric analysis demonstrated, similar to the canine data, that the SCD bound the more activated circulating neutrophils and monocytes. For neutrophils, the MFI of the cell surface marker for CD10 was used since it is more highly expressed on the cell surface as a neutrophil matures (47,48). A more mature neutrophil has a greater ability for proinflammatory activity (47). Accordingly, the SCD bound neutrophils had much higher MFIs for CD11b and CD10 compared to MFIs of the circulating neutrophils. These binding events were associated with a decline with SCD treatment time to lower levels of these two markers on circulating neutrophils and suggestive of immunomodulation of neutrophil activity.

For monocytes, the SCD bound monocytes once again had greater degrees of activation than circulating monocytes as reflected in the higher CD11b and CD14 MFIs of eluted cells. The impact of these SCD/monocyte interactions resulted in a less inflammatory state in the circulating pool of monocytes. This fact was reflected in the steady decline in the cell surface expression of HLA-DR on the intermediate (CD14+CD16+) subset of monocytes, a marker of monocyte proinflammatory activity (49). The cell surface expression of monocyte CD192 (CCR2) and CD197 (CCR7) were also measured in circulating and SCD bound monocytes. CCR2 is the receptor for monocyte chemotactic protein (MCP)-1 (46) and CCR7 is the receptor for chemokines CCL19 and CCD21. These chemokine ligand/receptor interactions critically modulate human monocyte adhesion and migration (47,48). The measurement of these surface markers demonstrated that SCD treatment resulted in a significant reduction in the CD197 MFI and an increase in CD192 MFI of the circulating monocyte pool. These changes may reflect important decreases in the release of CD197 expressing cells from monocyte stored pools and a decline in migration of CD192 expressing monocytes out of the circulation perhaps due to the decline of MCP-1 plasma levels with SCD treatment (Table 2, 46)

The kinetics of changes in the circulating pool of neutrophils were also evaluated. The amount of SCD eluted neutrophils after the first treatment was nearly 30% of the circulating pool and were comprised of highly activated leukocytes as reflected in the MFIs for CD11b and CD10. The subsequent analysis of membrane bound cells at day 3 and 5 demonstrated a sequestration of 3-5% of the circulating pool, values commonly seen in prior preclinical and clinical evaluations. This finding suggests an elevated number of activated neutrophils in this patient’s baseline CHF state. The distribution of the 3 subsets of circulating monocytes were also measured and demonstrated a shift after the first day of treatment from the classical (CD14++CD16-) toward the intermediate (CD14+CD16+) phenotype, suggesting release of a large proinflammatory pool of intermediate monocytes. This subset of cells eventually bound to the SCD as reflected in the high percentage of the eluted pool of monocytes being of the intermediate phenotype. This observation suggests that the SCD may deplete a reservoir of intermediate monocytes destined to migrate and maintain a M2 macrophage pool in myocardial tissue. The removal of this monocyte reservoir with SCD treatment may hinder the maintenance of the proinflammatory state within myocardium.

The effect of SCD treatment on plasma cytokine levels are noteworthy. All of the measured plasma cytokine levels decreased after 2 days of SCD treatment, demonstrating an immunomodulation of the hyperinflammatory state of this patient with longstanding HFrEF. The elevated IL-6 and MCP-1 concentrations were normalized. IL-6 is the prototypic proinflammatory cytokine while MCP-1 release from damaged myocardium in CHF promotes attraction and migration of monocytes into cardiac tissue (41,50). Since CD192 (CCR2) is the cell surface receptor to MCP-1, the reduction in MCP-1 levels correlates to the rise of CD192 expressing circulating monocytes observed in this patient after two days of SCD treatment.

This study is limited in that the data presented represents clinical experience with one patient. Recruitment continues for this study to gather safety and preliminary efficacy data.

In summary, the preclinical studies in a canine model of systolic CHF clearly demonstrated a SCD promoted durable improvement in myocardial contractility and cardiac performance over a 4-week period. This improvement was associated with SCD related immunomodulatory effect in these animals as determined by serum biomarkers as well as circulating neutrophil and monocyte phenotypes. The first in human, proof of concept translation of this SCD promoted improvement of cardiac performance was tested with a carefully designed clinical protocol with a reasonable benefit to risk profile. The first patient in this clinical evaluation demonstrated that SCD treatment quickly improved cardiac performance, as measured by elevations in multiple cardiac parameters including CPO and RVSVI, while maintaining renal function and net volume removal. These clinical outcomes were associated with SCD related immunomodulatory effecs as measure with plasma cytokine levels and leukocyte flow cytometry. Of importance, the first patient undergoing SCD treatment achieved the primary endpoint of this intervention of a successful LVAD implantation and discharge to home.

## Data Availability

All relevant data are within the manuscript and its Supporting Information files.

