## Supplemental Appendix for "Translation of Immunomodulatory Therapy to Treat Chronic Heart Failure: Preclinical Studies to First in Human"

**H. David Humes**

**The PDF file includes:**

Materials and Methods

Figs.S1 and S2

Tables S1

Supplemental References

### Materials and Methods

#### SCD formulation and Implementation.

The SCD is an extracorporeal cartridge containing polysulfone hollow fibers with blood flow directed to the side ports of the cartridge with closed end caps (Figure 1). Blood flow directed to the side ports enters the extra-capillary space of the device so that blood flows along the extraluminal side of the membranes with shear stress approaching capillary shear. This low shear stress allows activated leukocytes to adhere to the outer surface of the hollow fiber membranes while they traverse along the blood flow path. The SCD treatment requires regional citrate anticoagulation (RCA) to maintain at all times the iCa concentration in the blood circuit between 0.25 to 0.4 mmol/L This set up allows continuous cell processing of circulating neutrophils and monocytes to a less proinflammatory phenotype, thereby tempering the hyperinflammatory state of the effector cells of the innate immunologic system which are central to promoting the cytokine storm and subsequent tissue damage.

The SCD-1.0 was supplied by SeaStar Medical, This SCD has 1.0 $M^2$ blood contacting surface area. The SCD sustains the same blood flow settings of the CRRT circuit. To attain its immunomodulatory effect, the SCD requires a rigorously controlled low ionized calcium (iCa) concentration between 0.25 and 0.4 mM in the extracorporeal blood circuit achieved with regional citrate anticoagulation (RCA) and dialysate/replacement fluid with no calcium. As required in RCA protocol, replacement calcium is infused into the blood circuit exiting the SCD so that blood returning to the patient maintains a normal calcium level in the patient. A CRRT protocol was utilized with weight-based dialysate/ replacement fluid rates and fixed citrate-to-blood flow ratio, as opposed to a titration approach. Personalized dosing for calcium supplementation based on pre-calculated effluent calcium losses is also utilized to (a) achieve adequate circuit ionized calcium (iCa) (<0.4 mm/L) for any hematocrit level and hence plasma flow and (b) keep systemic citrate levels <2 mmol/L irrespective of body citrate metabolism and eliminate the risk of clinically significant hypocalcemia (1).

For this study, hypertonic ACDA citrate was administered into the circuit pre-hemofilter at 150-160 mL/hr. The subject was administered a calculated volume of 600 mL parenteral (in the form of D5W) free water during SCD treatment to convert the hypertonic sodium load from the ACDA into isotonic sodium gain. Finally, to remove the combined volume of ACDA and free water intake and the sodium load from the ACDA, isotonic to the patient ultrafiltration during SCD treatment, was 260 mL/hr for 6 hours to maintain no net volume change during SCD treatment. To maintain normocalcemia in the patient, CaCl2 was administered in a port just proximal to the blood return line targeting levels of 0.9 to 1.2 mM. Circuit and systemic iCa values were measured every 1-2 hours during SCD treatment.

The patient underwent therapy according to clinical protocol with SCD treatment for 6 hours daily for 6 days at which time a decision to proceed to LVAD implantation was made. Standard of practice (SOP) clinical laboratory values and 24-hour collections of urine for volume and analytes were obtained daily. With the presence of a Swan-Ganz catheter, cardiac and hemodynamic parameters were measured according to SOP protocol and recorded. Research blood samples for cytokines, biomarkers and cytometric analysis were obtained daily just prior to SCD treatment.

This low iCa is necessary to maintain anticoagulation within the circuit as well to provide the immunomodulation activity of the device. The low iCa environment within the extra-capillary space of the device promotes the selective binding of the most activated circulating neutrophils and monocytes in the blood due to the calcium requirement of cell surface molecules, including CD62L and CD11b/CD18 integrin, of these leukocytes to bind to surfaces (2,3). The more activated the cells, the more binding molecules are expressed on the cell surface, thereby increasing the density of these binding sites on the cell and more intense binding to the membranes. The leukocytes bind for minutes to hours on the membranes due to the low shear stress along the blood path within the SCD. Prolonged exposure of these cells in the low iCa environment within the SCD results in release of these cells back to the patient with a less proinflammatory phenotype resulting in a dampening of the overall activated state of the circulating population of neutrophils and monocytes. For neutrophils, recent data demonstrates that cell processing of neutrophils within the device results in the initiation of the apoptotic program of the bound neutrophils as has been reported in low iCa enviroments (4,5) with release back to the circulation where the normal clearance of apoptotic neutrophils occur via phagocytosis and digestion by macrophages within the bone marrow, spleen, and liver (6). For monocytes, cells released from the SCD back into the circulation shifts the circulating pool of monocytes to a less inflammatory phenotype (14). This continuous leukocyte processing results in the removal of the more proinflammatory leukocytes from the circulation and release of less inflammatory neutrophils and monocytes which are quickly removed from the circulation via normal homeostatic processes. This continuous process during a systemic inflammatory state results in an immunomodulatory effect to temper the excessive, dysregulated hyper-inflamed state and allows tissue repair and recovery.

**Materials and Methods for Flow Cytometry of Canine Samples**

Alexa Fluor ® 647 conjugated anti-canine CD14 antibodies, CD11b (unconjugated antibodies in supernatant, ABD SeroTec), and CFTM 488A conjugated anti-mouse IgG1(Sigma) were added to pre-chilled peripheral blood or elution collections. Red cells were lysed and leukocytes fixed by addition of Becton-Dickinson's FACS lysing solution. CD11b and CD14 surface expression was quantitated and reported as mean fluorescent intensity (MFI) with an Accuri C6 flow cytometer. Neutrophil activation was evaluated using a delayed apoptosis assay. Neutrophils isolated using a discontinuous Percoll gradient were resuspended in culture media and incubated for 24 hours, labelled with propidium iodide and FITC-Annexin-V (Abcam), then analyzed by flow cytometry (Accuri C6).

**Materials and Methods for Flow Cytometry and Cytokine Analysis of Clinical Samples**

Cytokines IL-1b, IL-6, IL-8, IL-10, TNFa, MCP-1, were analyzed with commercial ProcartaPlex Panels (ThermoFisher) and read using a Luminex by the University of Michigan's Cancer Center Immunology Cores. For cytometric analysis, erythrocytes were lysed immediately with ammonium chloride buffer, and cells washed in cold PBS with 0.2% EDTA and adjusted to $10^6$ cells per test. Live Dead Aqua (ThermoFisher L-34974) was added to exclude labeling artifacts caused by dead cells and followed by incubation with fluorochrome-conjugated antibodies. For monocyte phenotyping, antibodies targeting human CD91(clone: A2MR-a2), CD14(clone: TÜK4), CD16 (clone: 3G8), HLADR (clone: TU36), CC192 (clone: MEM-166) and CD197 (clone:150503) were used to determine changes in the circulating monocyte population. CD91 is useful as a pan monocyte marker (7). CD14 and CD16 allow for identification of classical, intermediate and non-classical phenotypes (8). HLADR is shed from the surface of monocyte as they become anergic to stimulatory signals (9) and assists with the identification of CD16+ intermediate (HLADRhigh) and non-classical (HLADRlow) monocytes (10). CD192 and CD197 correspond to the C-C motif chemokine receptors CCR2 and CCR7. CCR2 and CCR7 expression was increased on all monocyte subsets in CKD subjects compared with controls (11) and is useful in determining immunologic effects of therapy on the targeted patient population. Neutrophils were evaluated using anti-human CD66b (Clone:913542), CD10 (Clone: H10a), CD184 (CXCR4, Clone: 12G5), CD62L (Clone: DREG-56) and CD33 (CloneWM53). CD33 is expressed on immature NE (12) and CD10 is expressed on mature neutrophils and may provide a quantitative analysis of bone marrow neutrophil release (13,14). CD184 is expressed on aging NE marked for apoptosis (15) can be used as a measure of neutrophil aging and turnover(16). CD16 and CD62L have been used to define immunologically distinct subpopulations of neutrophils in response to inflammatory signals (17) and CD62L sheds upon binding of neutrophils to endothelium. Neutrophils [18,19] and monocytes (20,21) mobilize intracellular stores of CD11b to the cell surface as they become (primed) activated. Blood cells treated with a saturating amount of anti-human CD11b (clones ICRF44 and CBRM 1/5) detects all extracellular CD11b and the integrin only in the open/activated conformation respectively, allowing a real-time measurement of systemic acute neutrophil priming and activation (22).

Similar to CD11b, carcinoembryonic antigen related cell adhesion molecule 8, CD66b is translocated to the surface of neutrophils with stimulation from inflammatory mediators and released in soluble form after degranulation of secondary granules (23,24) It is also useful as a pan neutrophil marker.  SCD cytometric data was acquired with an Attune flow cytometer (ThermoFisher) and analyzed using FlowJo software. Panel compensations were auto-generated by Attune software using lot matched antibody labeled ARC Amine Beads or ABC compensation control beads (ThermoFisher) where appropriate. Antibody panels, manufacturer and channels used, are shown in Table S1 Figures S1 and S2 displays the gating hierarchy used for monocytes and neutrophils.

**Table S1. Flow Cytometry Reagents**

| Systemic Blood Panel 1: Monocytes | | | | |
|---|---|---|---|---|
| Target | Channel and label | Vendor | Catalog# | Clone |
| CD11b | BL1 FITC | BioRAD | MCA551F | ICRF44 |
| CD91 | BL2 PE | ThermoFisher | 12-0919-42 | A2MR-a2 |
| CD14 | BL3 PERCP Cy5.5* | BioRAD | MCA1568 | Tuk4 |
| | BL4 | | | |
| CD192 | RL1 Alexa Fluor® 647 | BD Biosciences | 558406 | 48607 |
| | RL2 | | | |
| CD16 | RL3 APCH7 | BD Biosciences | 560195 | 3G8 |
| CD197 | VL1 BD OptiBuild™421 | BioLegend | 353208 | Go43H7 |
| LIVE/DEAD | VL2 fixable Aqua | ThermoFisher | L34966 | NA |
| HLA-DR | VL3 BD OptiBuild™605, | BD Biosciences | 562845 | TU36 |
| | VL4 | | | |

* Antibody conjugated using LNK142PERCPCY5.5 Kit (Bio-Rad).

| Systemic Blood Panel 2: Neutrophils | | | | |
|---|---|---|---|---|
| Target | Channel | Vendor | Catalog# | Clone |
| CD11b | BL1 FITC | BioRAD | MCA551F | ICRF44 |
| CD11b activated | BL2 PE | BioLegend | 301406 | CBRM1/5 |

| | | | | |
|---|---|---|---|---|
| CD10 | BL3 PERCPCy5.5 | BD Biosciences | 563508 | HI10a |
| CD62L | BL4 PE CY7 | BD Biosciences | 565535 | DREG-56 |
| | RL1 | | | |
| CD33 | RL2 Alexa Fluor® 700 | BioLegend | 303436 | WM53 |
| CD16 | RL3 APCH7 | BD Biosciences | 560195 | 3G8 |
| CD66b | VL1 BD OptiBuild™421 | BD Biosciences | 562940 | G10F5 |
| LIVE/DEAD | VL2 fixable Aqua | ThermoFisher | L34966 | NA |
| | VL3 | | | |
| CD184 | VL4 BD OptiBuild™711 | BD Biosciences | 740799 | 12G5 |

B, R, and VL# refer to the Blue Red and Violet laser channels of the Attune Cytometer.

**Figure S1. Gating hierarchy for monocytes.** A. Singlets were selected in forward scatter height vs. forward scatter area. B. Live monocytes cells selected inside scatter vs. VL2 Live Dead Aqua. C. Monocytes were further selected using CD91 gated in side scatter area vs. BL2-CD91. Monocytes were then separated into classical, intermediate, and non-classical subset in scatter plot BL-3 CD14 vs, RL3 CD16 as shown in density plot. This graph is also shown as a Heat plots of HLA-DR, and was to confirm gating. Mean fluorescence intensity (MFI) for CD11b, CD14, CD16 CD197, CD192, and HLA-DR were calculated for the entire monocyte population and CD14/16 defined subgroups.

**Figure S2. Gating hierarchy for neutrophils.** A. Singlets were selected in forward scatter height vs. forward scatter area. B. Live cells selected in side scatter vs. VL2 Live Dead Aqua. C. Neutrophils gated in side scatter area vs. forward scatter area. D. Mean fluorescence intensity (MFI) for CD11b, CD10, CD16 CD33, CD62L, CD66b, and CD184.

Monocytes were then separated into classical, intermediate, and non-classical subset in scatter plot BL-3 CD14 vs, RL3 CD16 as shown in density plot. This graph is also shown as a Heat plots of HLA-DR, and was to confirm gating. Mean fluorescence intensity (MFI) for CD11b, CD14, CD16 CD197, CD192, and HLA-DR were calculated for the entire monocyte population and CD14/16 defined subgroups.
