## Supplementary figures and images for "Translation of Immunomodulatory Therapy to Treat Chronic Heart Failure: Preclinical Studies to First in Human"

### Fig. S1

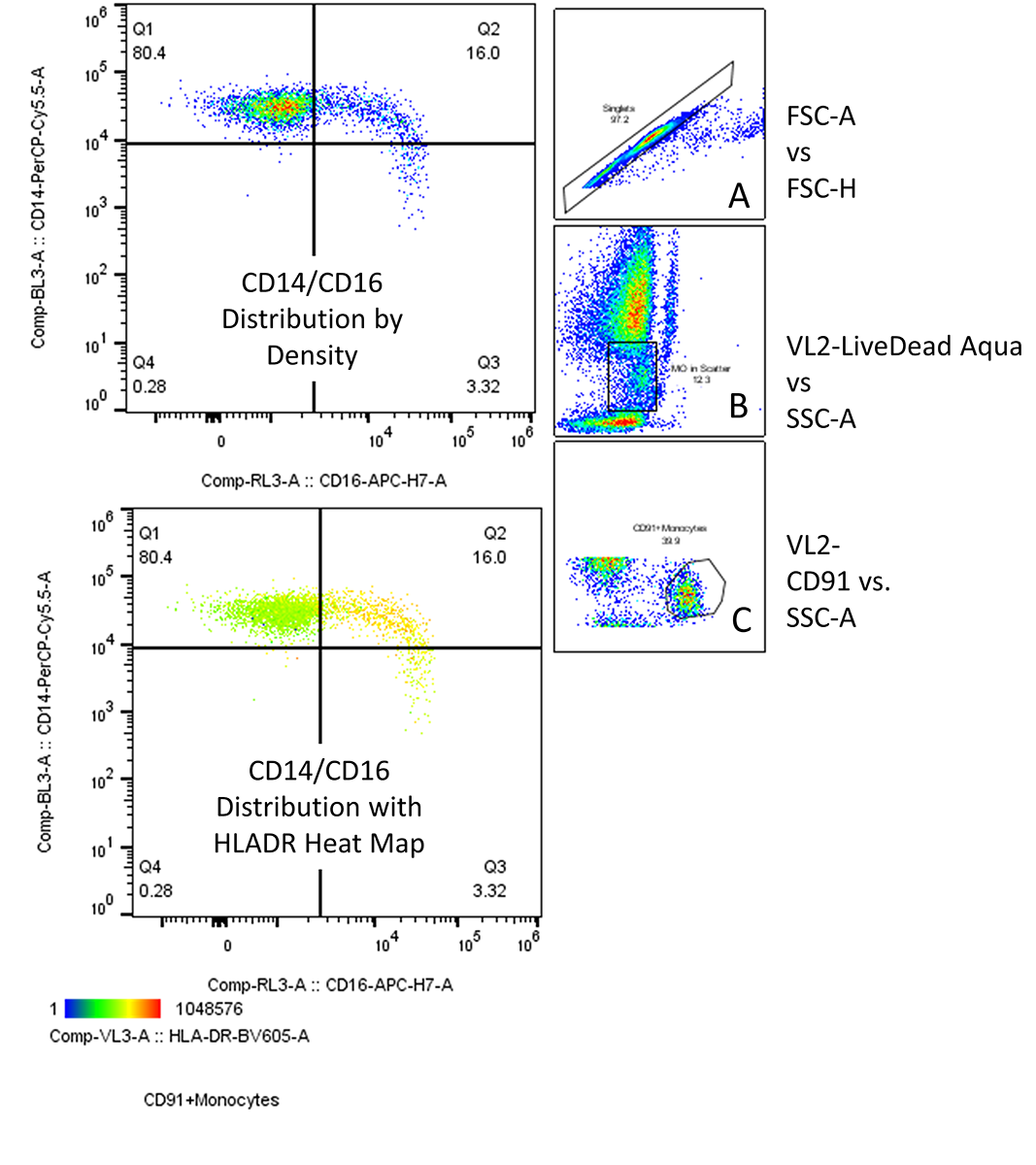

### Fig. S2

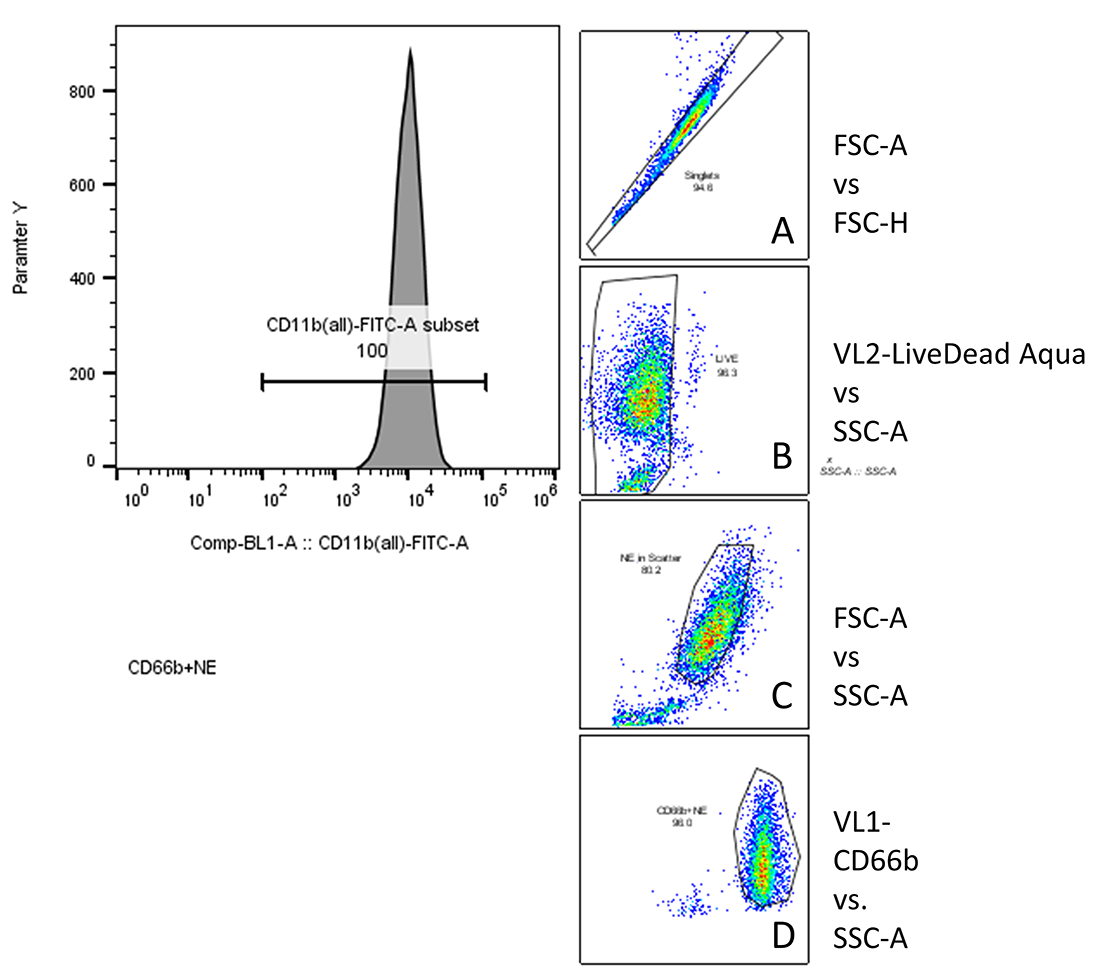
